# ANALYSIS OF FOUR DIFFERENT TRANSPORT AND PRESERVATION MEDIUM KITS FOR SARS-COV-2 DIAGNOSIS FROM NASOPHARYNGEAL SWAB BY REAL-TIME PCR: ADAPTING TO THE CONSTANTLY INCREASING DEMAND OF SAMPLING PROCESSING AND STOCK-OUTS DURING THE PANDEMIC

**DOI:** 10.1101/2021.07.13.21260473

**Authors:** Carlos Barrera-Avalos, Roberto Luraschi, Eva Vallejos-Vidal, Maximiliano Figueroa, Esteban Arenillas, Daniela Barría, Felipe Hernández, Carlos Mateluna, Javier Mena, Claudia Rioseco, Claudia Torrent, Claudio Vergara, Gaby Gutiérrez, Javiera Quiroz, Javiera Alarcón, Julio Cartagena, Javiera Cayunao, Andrea Mella, Álvaro Santibañez, Sebastián Tapia, Alejandro Undurraga, Deborah Vargas, Valentina Wong, Ailen Inostroza, Daniel Valdés, Mónica Imarai, Claudio Acuña-Castillo, Felipe E. Reyes-López, Ana María Sandino

## Abstract

The high demand for supplies during the COVID19-pandemic has generated several stock-out of material and essential reagents needed to meet the current high demand for diagnosis in the worldwide population. In this way, there is limited information regarding the performance of different virus transport medium (VTM) for nasopharyngeal swab sampling (NPS) aimed for SARS-CoV-2 detection. We compared the RT-qPCR amplification profile of four different commercial transport medium kits, including DNA/RNA Shield™, NAT™, VTM, and Phosphate-buffered saline (PBS) transport medium, for NPSs samples from Central Metropolitan Health Service, Santiago, Chile. The RT-qPCR showed a slight lower RNase P Cq value of the samples preserved and transported in DNA/RNA Shield™ compared to NAT medium. By contrast, a marked increase in the RNase P Cq value was registered in the samples transported with VTM compared to DNA/RNA Shield™ medium. For PBS-preserved NPS, the performance of two strategies were assessed due to the potential presence of any remaining active virus in the sample: (1) thermal inactivation; and (2) thermal inactivation treatment followed by RNA extraction. The heat inactivation showed a significantly lower Cq value for RNase P and viral ORF1ab Cq compared to the followed by RNA extraction. This study indicates that new medium alternatives could be used if supplies run out to diagnose COVID19.

## 1. INTRODUCTION

The severe acute respiratory syndrome coronavirus type 2 (SARS-CoV-2) has exposed the diagnostic research teams to several challenges in the process of his detection to control the COVID 19 pandemic. The current massive worldwide molecular method to detect SARS-CoV-2 genome in any sample is by RT-qPCR, a molecular diagnostic technique that identifies the genetic material of the virus from the upper respiratory tract, including oropharyngeal, nasopharyngeal or saliva samples [1–3]. Due to the large number of PCR testing, most labs had to deal with supply chain disruption, which became a limitation for COVID-19 testing due to lack of reagents [4]. Therefore, scientists are permanently exposed to frequent changes in viral transport medium (VTM), swabs, reagents for RNA extraction, diagnostic PCR kits for virus detection, among others. One of the critical steps for virus detection is the pre-analytical stage that involves collection, preserving, and transporting the sample to the clinical laboratory[5,6]. In terms of sampling collection, the Centers for Disease Control and Prevention (CDC) of the United States suggested the use of sterile synthetic fiber swabs with a plastic rod to avoid deterioration and, consequently, a possible compromise for the SARS-CoV-2 diagnosis. In addition, CDC also indicates that the sample should be transported in a suitable VTM for an efficient diagnosis of the COVID-19 [7]. On the other hand, The Food and Drug Administration (FDA) recommended the use of alternative VTM to counteract the constantly increasing demand in the transport and preservation of the viral sample [8]. Therefore, a wide spectrum of sampling, preservation and transport kits with different VTMs are currently produced to make up the stock-out in the pre-analytical stage. Despite the advantage that this apparently implies, it is necessary to determine the capacity of the different kits to preserve reproducibility in the SARS-CoV-2 diagnosis. Thus, the use of different preservation and transport kits must ensure the correct diagnostic for favoring the adequate applications of public health measures for controlling the pandemic.

The chemical composition of the transport medium can affect, for example, the integrity of the viral RNA. This has as repercussion a lower detection sensitivity on the PCR assay, thus bringing as consequence the report of samples with a false negative diagnostic [9,10]. Severe reports have studied VTM of a repertoire of kits in presence and absence of viral inactivators [11–14]From the lab technician safety perspective, the absence of viral inactivators on the VTM composition generates, as consequence, a prior processing step for viral inactivation. This process is made routinely by heat, for its safety manipulation and processing in type 2 biosafety laboratories [15]. In this study, we evaluated the performance of four commercial kits aimed for collection, preservation, and transport of nasopharyngeal swab samples (NPSs), for detection and diagnostic of SARS-CoV-2 by RT-qPCR. The solutions included in the analysis were based on DNA/RNA Shield, NAT medium (NAT), Virus Transport Medium (VTM; consisting in foetal bovine serum (FBS) added to a solution of Hanks’ salts (HS)), and phosphate-buffered saline (PBS) transport medium. These solutions were chosen because they were the only available in the Central Metropolitan Health Service in Chile. We observed differences in the amplification of the internal control (RNase P), probably associated to the different medium and preservation characteristics.

## 2. MATERIALS AND METHODS

### 2.1 Patient data collection for the first epidemic wave in Chile

This study includes the data for all patients received from the Central Metropolitan Health Service zone (Santiago, Chile) for SARS-CoV-2 diagnostic by RT-qPCR at the Laboratory of Virology (Center of Biotechnology and Aquaculture, University of Santiago of Chile). In total, 75,961 nasopharyngeal swab samples (NPSs) were received during the first epidemic wave, between April-September, 2020.

### 2.2 Samples

Nasopharyngeal swab samples (NPSs) of clinical patients that belong to the Central Metropolitan Health Service (Santiago of Chile) were included in the study. These samples are currently routinely processed in our role of laboratory of SARS-CoV-2 diagnostics and member of the University laboratories network developed in Chile for increasing the diagnostic capacity at national level. The swab samples were taken, preserved and transported using: (1) Genosur sampling and transport kit (catalog number: DM0001VR; Genosur LLC, NW) that contains an RNA stabilization buffer called DNA/RNA Shield (Zymo Research Corp. Irvine, CA) that immediately provoke virus inactivation potentially present in the sample. (2) IMPROVIRAL™ NAT medium (NAT) (catalog number: 550040; Improve Medical Instruments Co. Ltd; Guangzhou, China), specially designed to stabilize and preserve microbial and human nucleic acids (RNA/DNA) for prolonged time periods, based on guanidine salts. (3) SANLI-Medical Virus Transport Medium (VTM) (catalog number 780027; Liuyang Sanli Medical Technology Development Co.,Ltd; Liuyang, Changsha, China), consisted in foetal bovine serum (FBS) added to a solution of Hanks’ salts (HS) maintaining virus activity over a wider temperature range. This facilitates the extraction of nucleic acid for subsequent samples, as well as the isolation of the virus. (4) Phosphate-buffered saline (PBS) sterile solution (catalog number 634280; Winkler LTDA. Laboratorio de Microbiología, Santiago, Chile). The analysis was conducted in a pair ways comparison manner collecting NPS from the same patient with each kit analyzed. For the comparison between NPS samples preserved and transported in DNA/RNA Shield and NAT medium, samples from four volunteer patients were analyzed. Comparison between NPS samples preserved and transported in DNA/RNA Shield and VTM was made including eight NPS volunteer patients. For the comparison made between the two different processing strategies for samples transported in PBS solution, fourty-two NPS volunteer patients were assessed. All the samples arrived at the laboratory before the first 24 hours after the sampling collection taking care of the cold chain temperature (in the case of NPS-preserved VTM and PBS solutions).

### 2.3 Virus inactivation

Prior to RNA extraction, the samples transported in VTM and PBS solutions were inactivated by thermal shock at 56°C for 10 min in a thermoregulated bath, in a similar way to that previously reported [16].

### 2.4 Total RNA extraction

Total RNA extraction was carried out using the Total RNA purification kit (96 deep well plate format; Norgen Biotek Corp; Canada). Briefly, 250 µL of NPS from each patient was collected in a 1.5 ml tube and vortexed with 500 µL of lysis buffer (buffer RL: absolute ethanol; 1:1) during 1 min. Then, the solution was centrifuged at 14,000 x *g* for 5 min at room temperature. Subsequently, 700 µL of the lysate was transferred to a 96-filter plate and centrifuged at 1,690 x *g* for 6 min. The 96-filter plate was washed twice with 400 µL of wash solution A. After each wash the plate was centrifuged at 1690 x *g* for 4 min. Then, the plate was centrifuged at 1690 x *g* for 10 min to any volume trace. Finally, the total RNA was eluted using 70 µL of Elution solution A and centrifuged at 1690 x *g* for 7 min. The purified RNA was evaluated immediately by quantitative reverse transcription PCR (RT-qPCR).

### 2.5 SARS-CoV-2 detection by RT-qPCR

The detection of viral SARS-CoV-2 genome sequence was carried out using the ORF1ab probe (TaqMan™ 2019nCoV Assay Kit v1,Thermo Fisher Scientific, Boston, MA. Cat. No. A47532) using a one-step strategy. Positive internal control probes for ORF1ab and RNase P (TaqMan™ 2019-nCoV Control Kit v1; Thermo Fisher Scientific, Boston, MA. Cat. No. A47533) were included and assessed individually in the 96-well PCR plate. The polymerase from TaqMan™ Fast Virus 1-Step Master Mix (Applied Biosystems™, Santa Clara, CA. Cat. No. 44-444-36) was included in each reaction. Each reaction contained 5 µl of TaqMan™ Fast Virus 1-Step Master Mix 4X, 1 µl of ORF1ab assay 20X (FAM detector channel), 1 µl of RNase P assay 20X (HEX detector channel), 11 µl of nuclease free water, and 2 µl of extracted RNA sample. The amplification thermal conditions include the reverse transcription at 50 °C for 5 minutes, predenaturation at 95 °C for 20 seconds, followed by 40 cycles at 95 °C for 3 seconds and 60 °C for 30 seconds. All the RT-qPCR reactions were performed on the Agilent AriaMx Real-Time PCR System (Agilent Technologies, Part. Santa Clara, CA. No. G8830A). Quantification cycle (Cq) and relative fluorescence units (RFU) data were extracted from each NPS using the Agilent AriaMx software.

### 2.6 SARS-CoV-2 ORF1ab gene standard curve and detection limit for diagnosis

To determine the minimum detection limit for the RT-qPCR and the efficiency of amplification (E), a standard curve for ORF1ab amplification was generated. The assay includes five serial dilutions (from 10 to 1×10^5^ copies) of the positive ORF1ab gene internal control (TaqMan™ 2019-nCoV Control Kit v1; Thermo Fisher Scientific, Boston, MA. Cat. No. A47533). Reaction volumes and thermal conditions for amplification are the same above described. All the RT-qPCR reactions were performed on the Agilent AriaMx Real-Time PCR System.

### 2.7 Data representation and statistical analysis

GraphPad Prism 8 statistical software was used to analyze and plot the data obtained. From the total received samples from April to September 2020, the cumulative number of samples, frequency of age and gender of the patient (by month in total and positive samples) was graphed. For comparisons between different transport media, the Cq and RFU values were analyzed. A paired two-sided Student T-test was used to determine differences between two groups of samples preserved and transported in DNA/RNA Shield medium, NAT medium and VTM medium. In the case of the PBS-preserved nasopharyngeal swab (NPS) samples, an unpair two-sided Student T-test was used to determine differences between two processing strategies for by RT-qPCR. A *p*-value of < 0.05 was considered statistically significant.

### 2.8 Ethical Considerations

All the experimental procedures included in this study was authorized by the Ethical Committee of the University of Santiago of Chile (No. 226/2021) and the Scientific Ethical Committee of the Central Metropolitan Health Service, Ministry of Health, Government of Chile (No. 370/2021), and following the Chilean law in force. Data analysis used for this study was conducted only using the internal sample code numbers assigned at the moment to receive them for diagnostics purpose. Accordingly, the samples have been irreversible anonymized.

## 3. RESULTS

### 3.1 Sustained increased SARS-CoV-2 diagnosis demand on nasopharyngeal swab samples (NPSs) during the first epidemic wave

The Laboratory of Virology (Center of Biotechnology and Aquaculture, University of Santiago of Chile) have processed and analyzed until the end of September 2020, 75,961 nasopharyngeal swab samples (NPSs) for the diagnosis of SARS-Cov2 in the population belonging to the Central Metropolitan Health Service zone (Santiago, Chile) including 19 Ambulatory Health Centers and 1 Hospital (Fig 1A).

**Fig 1.**
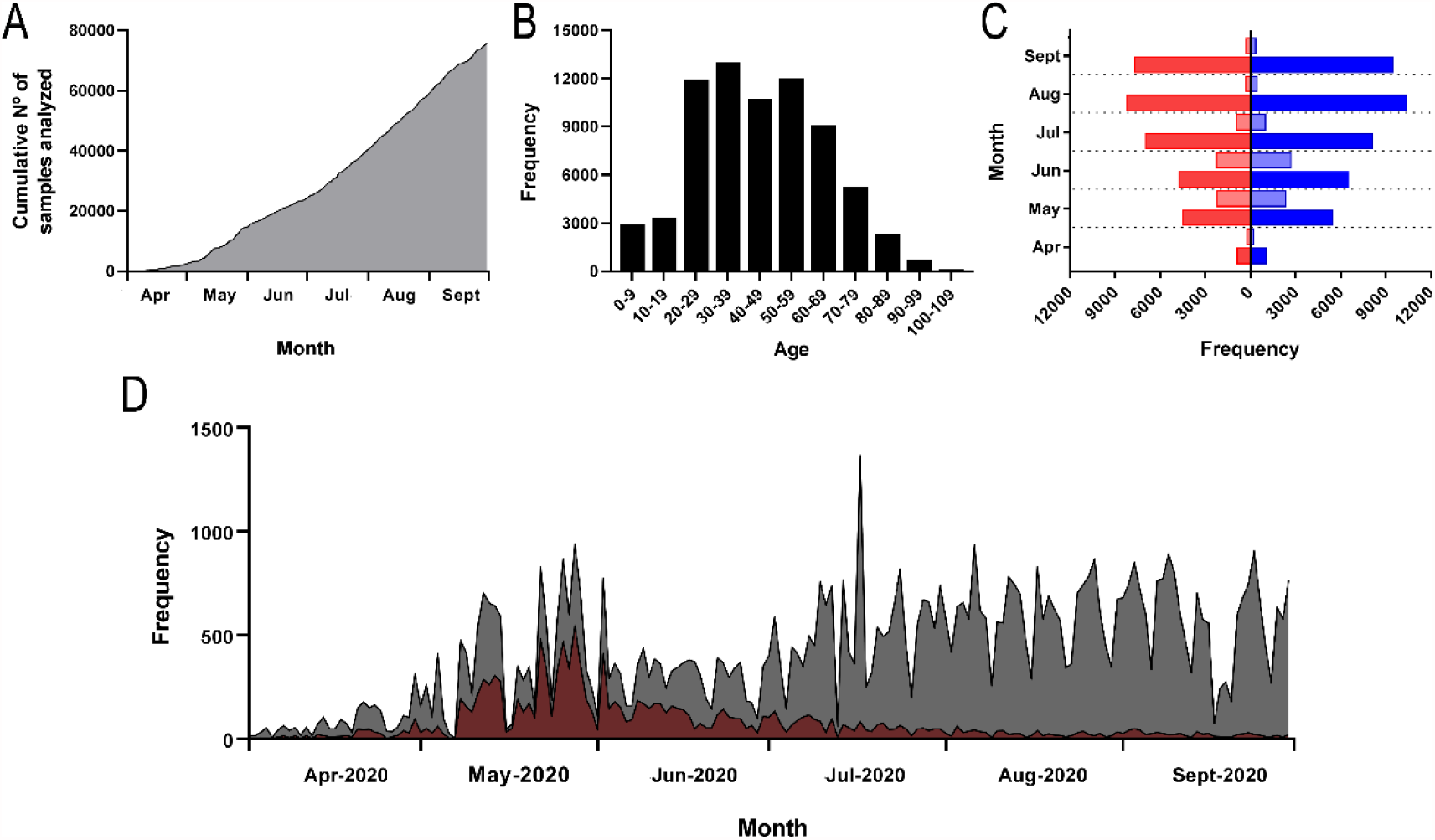
Diagnostic capacity of samples processed in the Virology laboratory (USACH, Chile) during the months of April and September 2020. (A) Cumulative number of samples analyzed. (B) Frecuency of age for nasopharyngeal swab samples analyzed for the diagnostic determination of SARS-CoV-2. (C) Distribution by month of total patients (male in solid red, women in solid blue) and positive patients (male in clear red, women in clear blue) distributed by gender. (D) Frequency of total (grey) and positive (dark red) daily samples during the period between April and September 2020.

The age spectrum of the patients analyzed ranged from newborns up to 105 years old. The age of the patients analyzed was mainly concentrated between 20 and 59 years (47,279 samples). Of these, the range between 30-39 years was the most analyzed age-interval group, reaching 12,915 patients. Then, it is followed by the intervals 50-59 years (11,876 samples), 20-29 years (11,843 samples), and 40-49 years (10,645 samples) (Fig 1B).

Between April and August, our laboratory registered a sustained increase in the capacity to analyze samples. Thus, our diagnostic laboratory went from 1,961 samples in April to processing 18,571 samples in August. A slight decrease in sample processing was recorded in September, reaching 17,158 processed samples. Regarding positivity, in April we registered 419 samples diagnosed as positive. A marked increase in the number of samples diagnosed as SARS-CoV-2 positive was observed in May (4,525), registering the peak of positivity in June with 4,947 samples. A greater positivity was observed for all months in females compared to males, reaching a peak of 2,692 samples in June (Fig 1C). Importantly, the high positivity was not associated with the number of samples analyzed. Indeed, on May 28, 936 samples were processed, of which 544 samples were diagnosed as positive. In contrast, on August 6 and September 24, 932 and 904 samples were processed, registering 41 and 20 positives, respectively (Fig 1D). Taken together, these results indicate that the positivity recorded in our team is related to the spread of SARS-CoV-2 infection in the population and not to the total number of samples processed.

### 3.2 The influence of preservation and transport solution kits upon the SARS-CoV-2 Cq values determined by RT-qPCR

The above-mentioned sustained increasing demand for supplies during the first pandemic wave generated several stock breaks of material and essential reagents to meet the current high demand for diagnosis in the worldwide population. For this reason, nasopharyngeal swab samples (NPSs) using different collection kits for transport and sampling preservation were analyzed. In our lab, most of the NPSs coming from the Central Metropolitan Health Service zone were preserved and transported routinely using the DNA/RNA Shield medium. However, because of the stock-out suffered due to the constant increase in the demand for diagnosis of SARS-CoV-2, we received NPSs from the Central Metropolitan Health Service samples in other two different transport mediums, including NAT and VTM-based transport kits. The evaluation of these NPSs kits with the internal control of total RNA extraction showed a slight lower RNase P Cq value of the samples preserved and transported in DNA/RNA Shield medium (16.36 ± 0.86) compared to the RNase P Cq vale for the samples preserved in NAT medium (18.59 ± 1.058) (Fig 2A). When comparing the relative fluorescence units (RFU) of samples conserved in DNA/RNA Shield and NAT medium (5,446 ± 493.5 and 5,124 ± 378.8 RFU, respectively), there was no significant difference (Fig 2B). No amplification for SARS-CoV-2 ORF1ab gene was observed in the four analyzed samples (Fig 2C).

**Fig 2.**
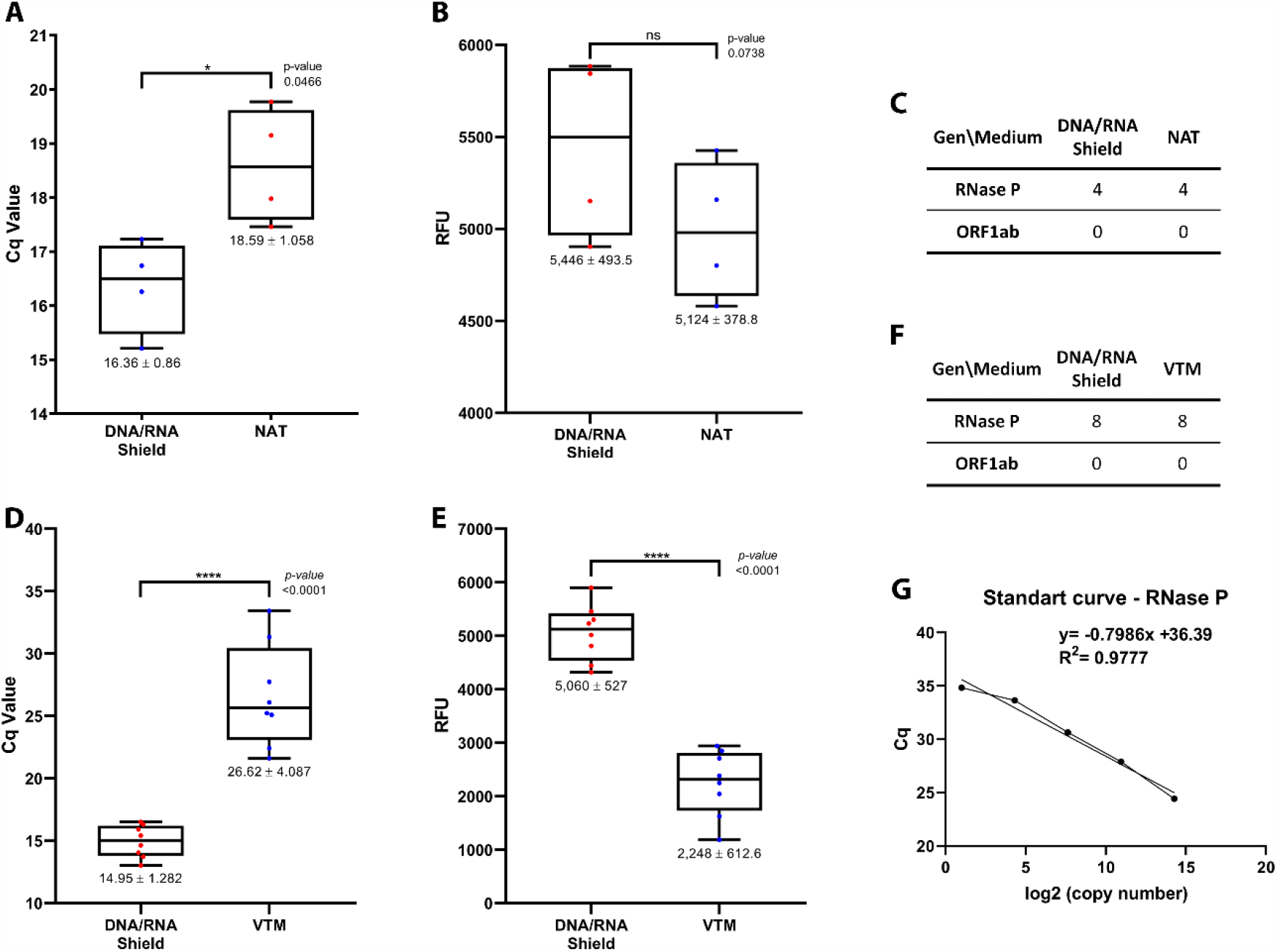
Determination of RNase P detection parameters for preserved nasopharyngeal swab (NPS) samples by RT-qPCR. (A) Cq value for paired data comparison between NPSs preserved and transported in DNA/RNA Shield and NAT medium (n= 4). (B) RFU (Relative fluorescence units) value comparison between NPSs preserved and transported in DNA/RNA Shield and NAT medium (n= 4). (C) Summary of the number of samples with RNase P amplification for NPSs preserved and transported in DNA/RNA Shield and NAT medium. (D) Cq value for data paired comparison between NPS samples preserved and transported in DNA/RNA Shield and VTM (n= 8). (E) RFU value for data paired comparison between NPS samples preserved and transported in DNA/RNA Shield and VTM. (E) Summary of the number of samples with internal control RNase P amplification for NPS samples preserved and transported in DNA/RNA Shield and VTM. (G) Standard curve for RNase P gene. Graph represent individual values and Q1, Q2 (median) and Q3 with whiskers at minimum and maximum values. For statistical t-test analysis, * *p*<0,05; ** *p*<0,01; *** *p*<0,001; **** *p*<0,0001.

On the other hand, a marked displacement in the value of Cq was appreciated in the samples transported with VTM for RNase P (26.62 ± 4.087), compared to the much slower RNase P Cq value obtained for the same samples preserved and transported in the DNA/RNA Shield medium (14.95 ± 1.282) (Fig 2D). Regarding the fluorescence intensity of the samples, those preserved in VTM medium showed a very low mean value (2,248 ± 612.6 RFU) compared to those samples from the same patients but preserved in DNA/RNA Shield medium (5,060 ± 527 RFU). (Fig 2E). No amplification for SARS-CoV-2 ORF1ab gene was observed in the eight analyzed samples (Fig 2F). All the analyzed samples registered a Cq value into the detection limit established for RNase P standard curve (Fig 2G).

In an extreme non-stock of preservation and transport kit for NPSs sampling, we started to receive them in 3 ml conic tube containing PBS solution. These PBS-preserved NPS samples were processed using two strategies: (1) thermal inactivation treatment at 56 °C for 10 minutes (for virus inactivation); and (2) the same thermal inactivation treatment followed by RNA extraction using Norgen kit. Then, the SARS-CoV-2 diagnosis was determined by RT-qPCR. Comparing the internal control RNase P Cq values obtained between the different sampling processing, the thermal inactivation showed a significantly lower value (21.18 ± 2.07) compared to the thermal inactivation followed by RNA extraction (27.62 ± 2.205) (Fig 3A). On the contrary, the relative fluorescence units for the RNase P internal control amplification showed higher RFU values (8,039 ± 955.1) for the thermal-inactivated samples than the thermal-inactivated samples followed by RNA extraction (5,756 ± 1,313) (Fig 3B). The same lower Cq value effect was observed for the viral ORF1ab gene detection in the thermal inactivated samples (25.73 ± 5.362) compared to the heat-treated and RNA extracted samples (30.27 ± 2.751) (Fig 3C). In the case of the viral ORF1ab gene, there are no significant differences between both sampling processing (Fig 3D). Importantly, with the thermal inactivation were diagnosed twenty-five SARS-CoV-2 positive samples; however, of them only nineteen samples were also diagnosed as SARS-CoV-2 positive (Fig 3E). All the SARS-CoV-2 positive-diagnosed samples fall into the range of detection for ORF1ab gene amplification (Fig 3F)

**Fig 3.**
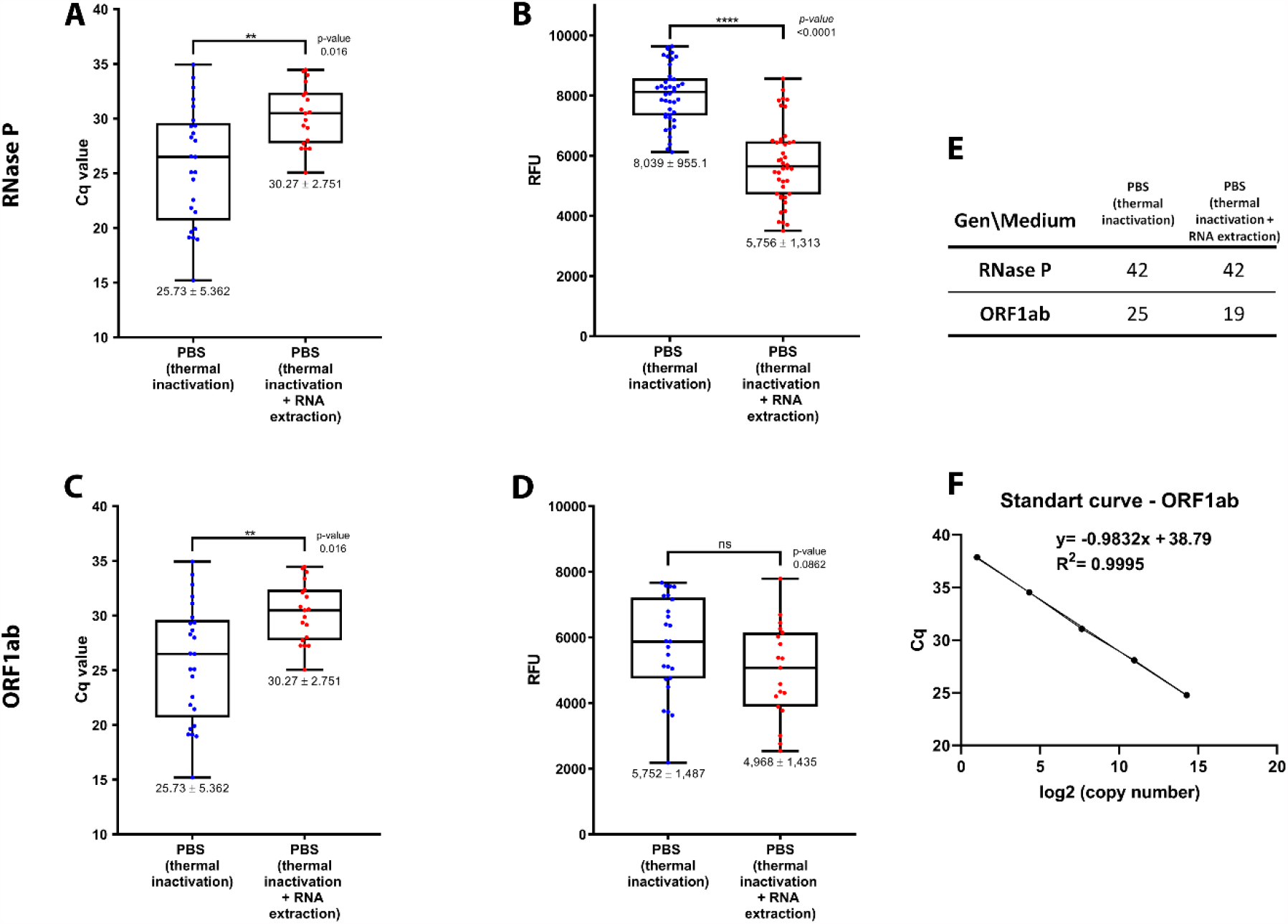
Comparison of two processing strategies for PBS-preserved nasopharyngeal swab (NPS) samples by RT-qPCR. RNase P (internal control) detection values for (A) Cq, and (B) RFU for thermal inactivation, and thermal inactivation followed by RNA extraction processing strategies. ORF1ab (Sars-Cov-2) detection values for (C) Cq and (D) RFU values for thermal inactivation, and thermal inactivation followed by RNA extraction. (E) Summary of the number of samples with RNase P amplification and ORF1ab amplification for both strategies assessed. (F) Standard curve for ORF1ab gene. For statistical Student t-test analysis, * *p*<0,05; ** *p*<0,01; *** *p*<0,001; **** *p*<0,0001. (n=42). RFU: Relative fluorescence units.

Collectively, the data for RNase P (cellular reference gene and used as an internal extraction control for human samples) suggest similar sample stability and preservation between DNA/RNA Shield and NAT medium. This antecedent indicates that the diagnostic result of the samples preserved with the VTM kit could be compromised, thus increasing the possibility of potential samples diagnosed as false negative. In the same way, in the PBS-preserved NPSs the thermal inactivation followed by RNA extraction could take place to the identification and report of false negative diagnostics.

## 4. DISCUSSION

The Real-time PCR (RT-qPCR) technique has been the gold standard and the most recommended method for the diagnosis for Coronavirus SARS-CoV-2 [17]. One of the critical stages for the diagnosis of this disease corresponds to the collection, preservation, and effective transport of the sample to the laboratory due to the nature of the RNA-genome of the virus. However, the high demand for supplies during the COVID19-pandemic has generated several stock breaks of material and essential reagents. As consequence, currently there is a wide repertoire of kits and for preserving and transporting the nasopharyngeal swab samples (NPSs) for the control of the pandemic. However, the variation in the detection signal and Cq value observed in our study strongly suggests that the transport and preservation medium should be considered for the detection of SARS-CoV-2. In this line, previous works have documented the efficacy of the use of different transport and preservation medium of the NPSs, either commercials [11–14] or in-house production [18–20], for the detection and diagnosis of Covid19. Accordingly, the Garnett et al 2020 [12] compared the performance of at least six swabs commonly found in primary health care settings for disease diagnosis; PurFlock Ultra, FLOQSwab, Puritan Pur-Wraps cotton tipped applicators, Puritan polyester tipped applicators, MedPro 6 “cotton tipped applicators, and HOLOGIC Aptima, which did not report significant differences in viral detection, indicating that several samples transport kit alternatives can be used if supplies run out. However, there are several other specimen transport and preservation kits used extensively for the diagnosis of the disease, which to date have not been studied or compared with each other. That is the case of the DNA/RNA Shield, NAT, VTM (FBS+HS), and PBS solution. In this study, the effect of different preservation and transport kits for NPSs was analyzed and compared for the massive diagnosis of SARS-Cov-2 by qPCR. We detected variations in the Cq value for the amplification of the internal cellular control, RNase P. These differences may be associated with the composition of the medium-in which the NPSs are preserved and transported. In the first place, in the case of the DNA/RNA Shield™ medium its composition stabilizes nucleic acids and biological samples at room temperature, in addition to completely inactivate viruses, bacteria and fungi. This agent has been widely used for sample collection and transport of various infectious agents [21–23]. Indeed, the group of Hamilton et al 2021[24] reported this reagent improves the detection of SARS-CoV-2 RNA in saliva samples, by detecting the Spike protein at low Cq cycles compared to samples preserved without DNA/RNA Shield™. Similarly, the group of Coryell et al 2021 [25] reported that DNA/RNA Shield™ gave the best results in terms of viral detection. Importantly, it also showed better stability compared to PBS solution after seven days of preservation, even after storage at −80°C. On the other hand, the viral transport medium (VTM) contains a solution of Hank’s salts supplemented with Fetal Bovine Serum. The absence of viral inactivator in the solution forces the NPSs inactivation by heat for its processing in class 2 biosafety laboratories. Thus, the sample handling associated to thermal inactivation followed by its processing by an RNA extraction step might affect the integrity of viral RNA. In fact, we observed a marked increase in the Cq value for RNaseP in VTM-preserved NPSs. Regrettably, there was no positive samples among those evaluated. Therefore, we cannot determine the impact of this higher Cq value for RNase P upon the SARS-CoV-2 diagnostic on NPSs.

This antecedent agrees with Graham et al, where a higher Cq values were observed in samples maintained in Hanks buffer solutions [26] compared to other types of medium, including DNA/RNA Shield™ solution. On the other hand, the NAT transport medium showed Cq and relative fluorescence values similar like those observed with DNA/RNA Shield™ solution. The NAT medium consists in an RNA stabilization solution with guanidine salts [27]. This is a strong chaotropic agent with the ability to deactivate viruses and preserve nucleic acids by disabling the action of nucleases and protein structures [28]. The use of this viral stabilizing and inactivating agent has been widely used in the study of several viruses [29,30] including SARS-CoV-2 for their detection and diagnosis during the pandemic [14,31,32]. For example, the Carvalho et al 2021 [33] analyzed NPSs kept in the medium with guanidine salts, obtaining Cq values like those reported in our current study. On the other hand, the PBS-preserved NPSs showed Cq values for RNase P higher than those obtained for DNA/RNA Shield™, NAT and even VTM solution in the case of a strategy following thermal inactivation and RNA extraction. This difference in the Cq value for the internal control could be associated to the absence of a stabilizing solution capable of preserving the quality of the viral RNA in transport kits containing only PBS medium, suggesting its heat inactivation prior manipulation. However, it has been supported the use of PBS as alternatives to VTM for SARS-CoV-2 testing [34,35] even preserving the sample at room temperature for 72 hours [12] or preserving the sample for a month at 4°C [36]. Anyway, our results agree with previous report that do not recommend the RT-qPCR analysis without a previous sample RNA extraction, due to the increase in Cq values in RNase P and viral gene [37]. However, the logistic difficulties for the primary healthcare centers to preserve the samples in PBS solution represent a serious disadvantage compared to those (i.e. DNA/RNA Shield™ and NAT media) that, in fact, do not require the maintaining of a cold chain temperature range from the sampling collection until processing. Furthermore, in the PBS-based transport kits there is a possibility the potential presence of any remaining still active viral particle; accordingly, prior inactivation is imperative for its safe handling in laboratory conditions with a class 2 safety level. The handling of active SARS-CoV-2 samples of respiratory viruses is recommended under strict protocols of type 3 biological safety laboratories [38,39]. Therefore, in our study the PBS-preserved samples were necessarily thermal inactivated as described in methodology section. The thermal-treated and RNA extracted samples showed a higher Cq value for the internal control amplification of RNase P, suggesting the process could affect the RNA integrity. In this way, the information about the RFU value is essential for the diagnosis, because samples with a low RFU are indicative of problems associated to sample degradation. Accordingly, in our results a lower RFU was also registered in the PBS-transported samples treated with thermal inactivation and subsequent RNA extraction. Undoubtedly, this fact could even compromise the diagnostic result. Indeed, a lower number of PBS-transported samples diagnosed as positives were identified compared to those samples that were just heat-treated. Thus, the thermal inactivation accompanied by an RNA extraction step compromise the SARS-CoV-2 diagnosis in NPS samples transported in PBS solution, increasing the chance for reporting false negatives diagnosis. Importantly, the thermal inactivation single-step treatment allows to obtain better results in less time-processing and lower cost associated with less use of consumables for diagnostics.

This study expands the knowledge about the performance of NPS transport solutions in pandemic circumstances and points out that the kits available on the market for collection, maintaining, and transporting samples and the modulation of Cq value range obtained for the internal control and SARS-CoV-2 detection. In this way, the displacement of Cq towards higher values may compromise the sensitivity of the PCR kit for the diagnosis of SARS-CoV-2. This increases the possibility to get potential false negative results. On the other hand, our results convincingly indicate that media containing viral inactivators such as DNA/RNA Shield ™ and guanidine salts give better parameters, also considering that it avoids viral inactivation by heat and disintegration of the RNA sample, being easier its handling and processing in type 2 biosafety laboratories. Future analyzes should include, for example, an evaluation of the quality preservation of the sample through the days, and at different range of temperatures. This study broadens the knowledge regarding the data obtained from NPS transport kits available in the market facing a stock out event in SARS-CoV-2 pandemic circumstances. These results also serve as evidence for the application of these transport media in the massive diagnosis of new emerging infectious diseases, which facilitates the choice of the most viable sample transport kit. This is particularly relevant in the context of respiratory viruses spreading to countries where the laboratory available massive infrastructure allows the handling and processing of samples only in a class 2 biosafety condition.

## 5. CONCLUSION

Collectively, our results indicate that the DNA/RNA shield solution and the NAT transport medium kit show a better performance in terms of Cq and RFU in the RNase P internal control and compared to the VTM kit. The heat inactivation of the viral sample preserved in PBS shows better results, in terms of RFU and Cq value, for the RNase P internal control and the SARS-CoV-2 ORF1ab gene amplification in the RT-qPCR analysis when its performance was compared to the thermal inactivation followed by RNA extraction step. Thus, the thermal inactivation single-step treatment allows to obtain better results in less time-processing and lower cost associated with less use of consumables for diagnostics, ensuring the operator safety in the handling and manipulation in biosafety class 2 laboratories of samples potentially in presence of SARS-CoV-2

## Data Availability

All relevant data are within the manuscript. Some anonymised data may be requested upon formal and justified request to the corresponding author

## Notes

### Competing Interest Statement

The authors have declared no competing interest.

### Funding Statement

A.S. 21180465. National Research and Development Agency. https://www.anid.cl/.

A.M.S. COVID1038. National Research and Development Agency. https://www.anid.cl/.

M.I. 1201664. National Fund for Scientific and Technological Development. https://www.anid.cl/.

F.E.R-L. 1211841. National Fund for Scientific and Technological Development. https://www.anid.cl/.

C.A-C. 021943AC. Directorate of Scientific and Technological Research.

https://www.vridei.usach.cl/direccion-de-investigacion-cientifica-y-tecnologica-0.

EV-V. EQM200016. Fondo de Equipamiento Cientifico y Tecnologico. https://www.anid.cl/.

### Author Declarations

All the experimental procedures included in this study was authorized by the Ethical Committee of the University of Santiago of Chile (No. 226/2021) and the Scientific Ethical Committee of the Central Metropolitan Health Service, Ministry of Health, Government of Chile (No. 370/2021), and following the Chilean law in force.

